# Predicting Hospital Utilization and Inpatient Mortality of Patients Tested for COVID-19

**DOI:** 10.1101/2020.12.04.20244137

**Authors:** Connor Davis, Michael Gao, Marshall Nichols, Ricardo Henao

## Abstract

Using structured elements from Electronic Health Records (EHR), we seek to: *i*) build predictive models to stratify patients tested for COVID-19 by their likelihood for hospitalization, ICU admission, mechanical ventilation and inpatient mortality, and *ii*) identify the most important EHR-based features driving the predictions. We leveraged EHR data from the Duke University Health System tested for COVID-19 or hospitalized between March 11, 2020 and August 24, 2020, to build models to predict hospital admissions within 4 weeks. Models were also created for ICU admissions, need for mechanical ventilation and mortality following admission. Models were developed on a cohort of 86,355 patients with 112,392 outpatient COVID-19 tests or any-cause hospital admissions between March 11, 2020 and June 4, 2020. The four models considered resulted in AUROC=0.838 (CI: 0.832-0.844) and AP=0.272 (CI: 0.260-0.287) for hospital admissions, AUROC=0.847 (CI: 0.839-855) and AP=0.585 (CI: 0.565-0.603) for ICU admissions, AUROC=0.858 (CI: 0.846-0.871) and AP=0.434 (CI: 0.403-0.467) for mechanical ventilation, and AUROC=0.0.856 (CI: 0.842-0.872) and AP=0.243 (CI: 0.205-0.282) for inpatient mortality. Patient history abstracted from the EHR has the potential for being used to stratify patients tested for COVID-19 in terms of utilization and mortality. The dominant EHR features for hospital admissions and inpatient outcomes are different. For the former, age, social indicators and previous utilization are the most important predictive features. For the latter, age and physiological summaries (pulse and blood pressure) are the main drivers.

## 1 Introduction

SARS-CoV-2, the etiological agent of the disease known as COVID-19, has been responsible for more than 38 million infections and over 1 million deaths globally as of October 14, 2020^1^. By the same date, in the United States (US) alone, the number of confirmed infections and deaths are estimated to be over 7.9 million and 216,000, respectively^1^. The COVID-19 pandemic has caused unprecedented stress on health systems worldwide that have resulted in a generalized population health emergency^2^. Challenges associated with the current pandemic manifest in many cases as prohibitive utilization of inpatient and Intensive Care Unit (ICU) facilities, ventilation equipment, as well as prolonged hospital stays that amplify the excess of utilization^3^.

It is well understood that SARS-CoV-2 primarily affects the respiratory system^4,5^. However, complications on other systems also contribute to increased hospital resource utilization and poor outcomes; including mortality^2^. The reported heterogeneity of the outcomes cascading from SARS-CoV-2 infections^4,5^, makes the accurate prediction of outcomes across the wide range of clinical presentations, a key consideration for the resourcing and management of patients with COVID-19, specially, in large health systems and urban areas with the need to triage large volumes of patients.

Electronic Health Records (EHR) though routinely used for clinical practice, host a vast amount of information about the medical history of patients that has been proven useful for predictive purposes in many scenarios^6–9^. Consequently, it also has the potential of being useful in the context of COVID-19. Unfortunately, data access limitations^10^, particularly, to the datasets from relevant COVID-19 cohorts may in some cases hinder progress, specially, in time-sensitive scenarios such as a global pandemic. Nevertheless, methods for predictive models based on both statistical and machine learning approaches have been explored, to address different predictive tasks associated with COVID-19, *e.g.*, infection rates and mortality forecasting, outbreak detection, (rapid) diagnosis from medical images or molecular markers, and prediction of outcomes (prognosis)^11–17^.

From the perspective of outcomes prediction, a shortcoming of existing approaches is their focus on mortality, which forgoes key utilization indicators such as hospital admissions, ICU admissions and the need for mechanical ventilation. In this work, we build models to predict inpatient mortality, as well as these key utilization metrics, on a cohort of patients tested for COVID-19 or hospitalized within the Duke University Health System (DUHS), Durham, NC, USA. In doing so, we account for the key transitions patients may experience in their medical-care journey after being tested, and some diagnosed, with SARS-Cov-2 (see Figure 1). Further, we also aim to identified the most important predictors from these models, as a means to provide some intuition into the patient characteristics driving the model predictions. Additionally, to encourage the development of clinical decision support systems conducive to more accurate risk stratification tools for patients, which may result in more effective hospital resource utilization and more appropriate care for those at higher risk of poor outcomes, specially, mortality.

**Figure 1.**
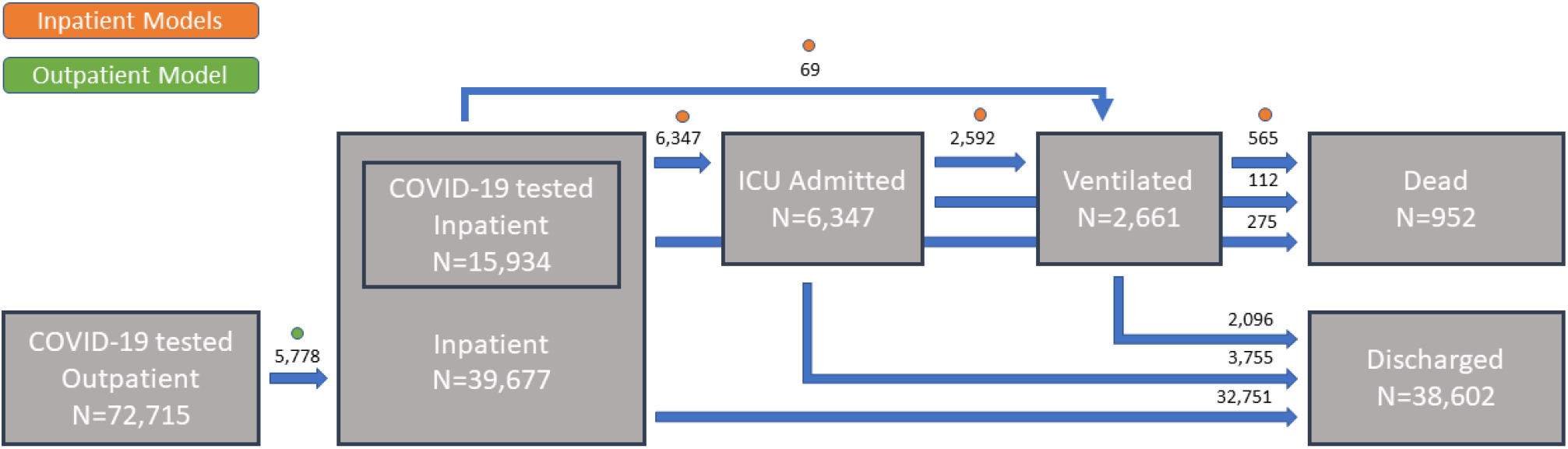
Patient flow summary. Patients can be thought as being in one of six states: outpatient, inpatient (admitted to the hospital), ICU admitted, ventilated, dead or discharged. Patients can transition between some of these states. In every state and possible transition, encounter counts for the DUHS cohort are provided. Dots indicate the state transitions for which the proposed models are will make predictions.

## 2 Materials and Methods

### Cohort

Two cohorts were identified for the purpose of predicting COVID-19 utilization and mortality outcomes. The *outpatient cohort* is defined as all adult patients with an encounter at any DUHS hospital facility who were not admitted to inpatient at the index encounter and who received a COVID-19 test until August 24, 2020. Because the first recorded COVID-19 test was administered on March 11, 2020, the cohort includes only patients after this date. All PCR-based clinical laboratory tests for SARS-CoV-2 that are here considered as COVID-19 tests were identified with the help of expert clinicians. Please refer to Appendix 8 for the details. The second cohort, the *inpatient cohort*, consists of all adult inpatient encounters at DUHS hospitals (Duke University Hospital, Duke Regional Hospital, and Duke Raleigh Hospital) admitted between January 1, 2020 and August 24, 2020. For both cohorts, adult patients are defined as 18 years and older. There are no other exclusions for either cohort.

### Data and Processing

Data were pulled from EPIC Clarity via the data pipeline developed by the Duke Institute for Health Innovation (DIHI). Elements pulled for both cohorts include age, encounter-specific items (time since last inpatient encounter, number of encounters at Duke Hospitals within the past year), procedures within the past year, and the active patient problem list within the past year; all relative to the index encounter, *e.g.*, a COVID-19 test in an outpatient setting or an inpatient encounter, regardless of COVID-19 testing, in the covered time frame (March 11, 2020 through August 24, 2020). Further, relevant vital signs recorded within the first hour of the encounter for the inpatient cohort and recorded prior to the COVID-19 test for the outpatient cohort were extracted. Data pulled only for the outpatient cohort includes sex, race, allergies, and social indicators (alcohol and tobacco use). Data pulled only for the inpatient cohort includes ICD-10 diagnoses within the past year, chief complaint at the time of inpatient presentation, and laboratory values, orders placed, and the Medication Administration Record (MAR) for the first hour of the index encounter.

Problem list and ICD-10 diagnosis data were grouped using the ICD-10 to single-category CCS crosswalk, and prior procedures were similarly grouped using the CPT to single-category CCS crosswalk^18^. The ICD-10 diagnosis data for the inpatient cohort were further divided into diagnoses assigned within the 90 days prior to the index encounter and those assigned within the past year but not within the past 90 days. For chief complaint data, only the most common 90% of complaint names were used; all other complaints were grouped into a single category. Other EHR data, including vitals, laboratory results (values), orders placed, and the MAR were grouped using crosswalks developed by DIHI with clinical collaboration. For data with numerical values, *e.g.*, laboratory results, an *missingness* indicator variable was also created to denote whether the item was recorded for a given patient encounter. See Appendix 8 for a list of all features used in the modeling.

The cohorts were divided into train, validation, and test sets. The test set was defined chronologically consistent with practical implementations, *i.e.*, as all encounters after a predefined date. The inpatient cohort test set includes encounters after June 4, 2020, and the outpatient cohort test set includes encounters after July 4, 2020. Choosing different dates for the outpatient and inpatient cohorts allowed for a more even partition of data between the train, validation and test sets. The train and validation sets consist of a random 70% and 30% split of all encounter data, respectively, prior to the starting dates for the test set. Prior to modeling, outliers from numerical features were removed by setting any values beyond the 99 and 1 percentiles to missing, and subsequently standardized (zero mean and unit standard deviation) using the means and standard deviations from the training set. Lastly, missing values were imputed to zero (mean values).

### Outcomes

The outcome of interest for the outpatient cohort is defined as inpatient admissions to a DUHS hospital within 4 weeks of the index encounter. For the inpatient cohort, ICU admission, mechanical ventilation, and inpatient mortality were considered. Units considered ICUs at DUHS hospitals were identified, and patients for which care was provided in one of these units during their inpatient stay were marked as ICU admits. Ventilation was identified from the EHR flowsheets. Specifically, patients that were assigned a ventilation order at any time during their encounter were marked as ventilated. Patients that died during the inpatient stay were identified as having a recorded date and time of death prior to, or concurrent to, the recorded discharge date and time.

### Statistical Models

Separate models were trained for predicting admission within 4 weeks of an outpatient COVID-19 test (outpatient cohort), and ICU admission, ventilation, and mortality (inpatient cohort). For each of these, three classification approaches were considered, one regression model and two gradient boosted models, namely, logistic regression, XGBoost^19^, and LightGBM^20^. After building simple models with each of the three approaches using the default hyperparameters on the train set, and estimating performance on the validation set, only the modeling approach with the best performance was used for further analysis. See Fig. S1 in the Supplement for validation performance of all modeling approaches, from which is clear that LightGBM performed the best. Consequently, all hyperparameter tuning (on the validation set) and final model training and testing was carried out with a LightGBM classifier.

Hyperparameter tuning was performed by using the RandomizedSearchCV function in Sklearn (Phyton package)^21^ on the training and validation sets. For this step, models were trained using 5-fold cross-validation, and early stopping was set to 10 rounds. See Table S1 in the Supplement for the range of values considered for each of the four models trained. Once optimal the hyperparameters were obtained, final models were trained with these hyperparameters on the combined train/validation set and were tested on the withheld test set.

Once the final models were built, the importance of each feature to the model predictions was determined using the importance attribute estimator from LightGBM^20^. Feature importance values were normalized so the most important feature would have an importance of 1. Features were then ranked in order of decreasing importance. The top-10 most important features are reported in the results.

### Performance Evaluation

Performance of the initial LightGBM classifier models, built only on the training sets, was determined by calculating the Receiver Operating Characteristic (ROC)^22^ and Precision-Recall Curves (PRC)^23^, as well as the Area Under the ROC curve (AUROC) and the Average Precision (AP) score. Classification thresholds for each model were selected using the Youden statistic^24^. For each model, the True Positive Rate (TPR), True Negative Rate (TNR), False Positive Rate (FPR), False Negative Rate (FNR), Positive Predictive Value (PPV), Negative Predictive Value (NPV), and accuracy were calculated using the threshold, *τ*, selected by Youden’s method.

The same metrics were calculated for the final models, built on the combined train and validation sets, using the withheld test sets. Specifically, encounters from July 4, 2020 to August 24, 2020 for the outpatient cohort and encounters from June 4, 2020 to August 24, 2020 for the inpatient cohort. The resulting ROC and PRC curves were compared against those for the validation set. The thresholds estimated via the Youden statistic for the validation sets were used to calculate TPR, TNR, FPR, FNR, PPV, NPV, and accuracy for the final models on their respective test sets. For all metrics, 95% Confidence Intervals (CI) were estimated via bootstrapping.

## 3 Results

### Cohort Characteristics

The characteristics of the cohort considered in the experiments below are presented in Tables 1 and 2 for outpatient in inpatient cohorts, respectively. For Age we report medians and Inter-Quantile Ranges (IQR) grouped by COVID-19 positive and negative patients. For the other cohort characteristics which are discrete, we report counts and proportions grouped similarly as Age. For the inpatient cohort in Table 1 we report the same characteristics in addition to Length of Stay (LOS) in days, for patients admitted to a DUHS hospital between March 11, 2020 and August 24, 2020, regardless of COVID-19 testing. In this case, the patients not tested for COVID-19 are denoted as *unknown* in Table 1.

**Table 1.**
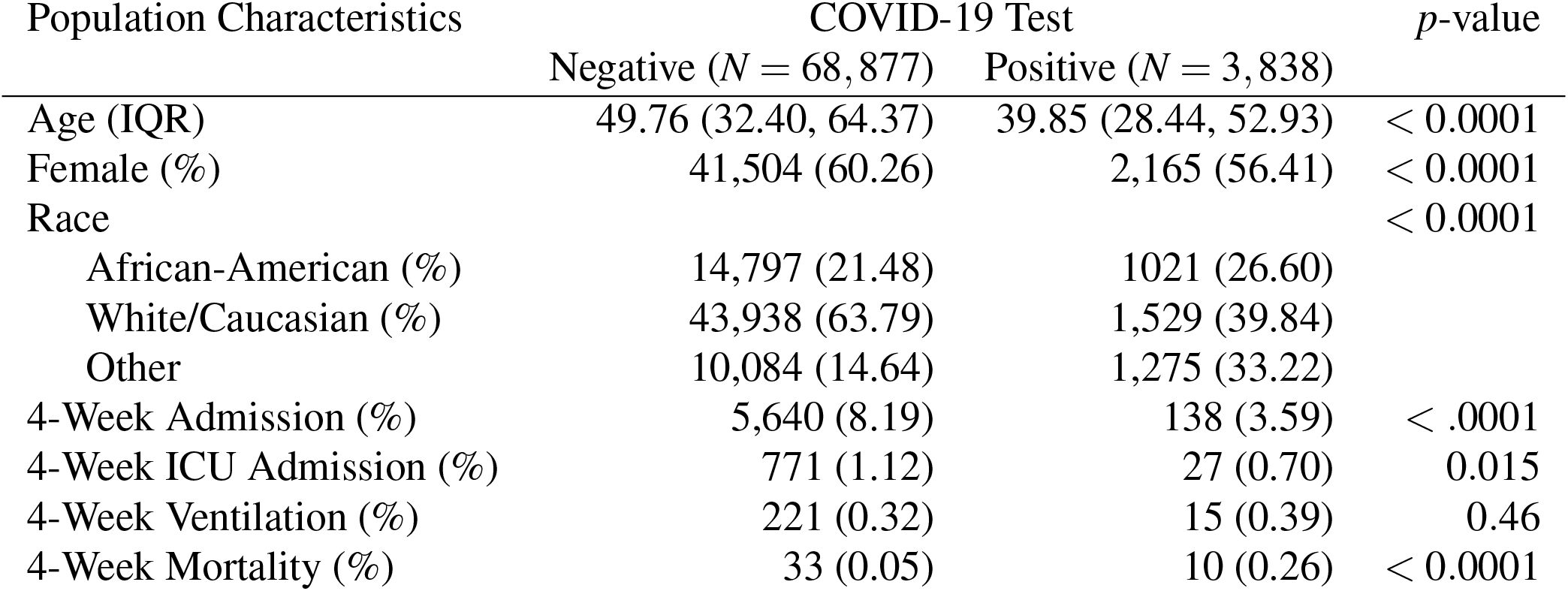
Outpatient cohort characteristics for patient encounters where a COVID-19 test was performed in an outpatient setting. ICU admissions, ventilation and mortality are for inpatient encounters within 4 weeks of COVID-19 testing. *p*-values were obtained using Wilcoxon rank sum test for Age and *χ*^2^ for all other comparisons.

| Population Characteristics | COVID-19 Test |  | *p*-value |
| --- | --- | --- | --- |
|  | Negative (*N* = 68,877) | Positive (*N* = 3,838) |  |
| Age (IQR) | 49.76 (32.40, 64.37) | 39.85 (28.44, 52.93) | < 0.0001 |
| Female (%) | 41,504 (60.26) | 2,165 (56.41) | < 0.0001 |
| Race |  |  | < 0.0001 |
| African-American (%) | 14,797 (21.48) | 1021 (26.60) |  |
| White/Caucasian (%) | 43,938 (63.79) | 1,529 (39.84) |  |
| Other | 10,084 (14.64) | 1,275 (33.22) |  |
| 4-Week Admission (%) | 5,640 (8.19) | 138 (3.59) | < .0001 |
| 4-Week ICU Admission (%) | 771 (1.12) | 27 (0.70) | 0.015 |
| 4-Week Ventilation (%) | 221 (0.32) | 15 (0.39) | 0.46 |
| 4-Week Mortality (%) | 33 (0.05) | 10 (0.26) | < 0.0001 |

**Table 2.** Inpatient cohort characteristics for patient encounters in a hospitalization setting. LOS is short for length of stay (in days). *p*-values were obtained using a Wilcoxon rank sum test for Age and LOS and *χ*^2^ for all other comparisons.

| Population Characteristics | COVID-19 Test | | $p$-value |
| --- | --- | --- | --- |
| | Negative or Unknown ($N = 38,756$) | Positive ($N = 921$) | |
| Age (IQR) | 61.14 (42.85, 72.54) | 60.02 (45.09, 71.85) | 0.6 |
| Female (%) | 21,280 (54.91) | 442 (47.99) | < 0.0001 |
| Race |  |  | < 0.0001 |
| African-American (%) | 12,532 (32.33) | 324 (35.18) |  |
| White/Caucasian (%) | 22,082 (56.98) | 282 (30.62) |  |
| Other | 4,141 (10.68) | 315 (34.20) |  |
| ICU Admission (%) | 6,276 (16.19) | 306 (33.22) | < 0.0001 |
| Ventilation (%) | 2,488 (6.42) | 173 (18.78) | < 0.0001 |
| Mortality (%) | 956 (2.47) | 119 (12.92) | < 0.0001 |
| LOS days (IQR) | 3.65 (2.15, 6.71) | 6.18 (3.22, 12.20) | < 0.0001 |

### Model Validation

The complete cohort of patients summarized in Tables 1 and 2 can be visually represented as the patient flow in Figure 1. From the total 112,932 encounters corresponding to 86,355 patients, 72,715 were tested for COVID-19 in an outpatient setting, 15,934 were tested for COVID-19 in an inpatient setting and 39,677 patients were hospitalized in the time frame considered (March 11, 2020 through August 24, 2020), regardless of COVID-19 testing. In fact, during this period 19,170 encounters correspond to inpatient events without concurrent or prior COVID-19 testing. From all hospitalization encounters, 38,602 (97%) resulted in hospital discharge, 6,347 (16%) were ICU admissions, 2,661 (7%) required mechanical ventilation and 952 (2%) died in an inpatient setting. Further, only 5,6778 (8%) outpatient encounters with a concurrent COVID-19 test resulted in inpatient admissions within 4 weeks of testing. Patient counts for detailed transitions are presented in Figure 1.

We consider two models, an outpatient model to predict hospital admissions within 4 weeks for patients tested for COVID-19 in an outpatient setting, and an inpatient model to predict ICU admission, need for mechanical ventilation and inpatient mortality. Models were developed for outpatient testing from March 11, 2020 through July 4, 2020, and for inpatient encounters from March 11, 2020 through June 24, 2020. The validation sets consists of a 30% random sample of these development sets. The test set consists of all encounters for patients with outpatient and inpatient encounters after July 4, 2020 and June 4, 2020, respectively.

Figure 2 shows graphical, threshold-free, performance characteristics on the validation and test sets, and the four models considered for the patient flow in Figure 1, namely, hospital admissions for patients tested with COVID-19 in an outpatient setting, ICU admission, mechanical ventilation and inpatient mortality. Specifically, we present the Receiving Operating Characteristic (ROC) and the Precision-Recall curve (PRC), along with the Area Under the ROC (AUROC) and the average precision (AP) for the PRC. ROC and PRC results on the test set grouped by COVID-19 testing status (positive, negative and not tested) are shown in Fig. S2 in the Supplement. Summaries of the ROC and PRC, *i.e.*, AUROC and AP, are also presented in Table 3 for both validation and test sets; including 95% Confidence Intervals (CIs) estimated via bootstrapping.

**Table 3.** Performance summary metrics for all models on the validation and test sets. We report AUROC (area under the ROC) and AP (average precision of the PRC). Figures in parentheses are 95% CIs estimated via bootstrapping.

| Model | Validation Set |  | Test Set |  |
| --- | --- | --- | --- | --- |
|  | AUROC | AP | AUROC | AP |
| Admission | 0.831<sub>(0.820,0.842)</sub> | 0.336<sub>(0.313,0.365)</sub> | 0.838<sub>(0.832,0.844)</sub> | 0.272<sub>(0.260,0.287)</sub> |
| ICU Admission | 0.846<sub>(0.837,0.856)</sub> | 0.625<sub>(0.604,0.649)</sub> | 0.847<sub>(0.839,0.855)</sub> | 0.585<sub>(0.565,0.603)</sub> |
| Ventilation | 0.845<sub>(0.828,0.863)</sub> | 0.330<sub>(0.287,0.373)</sub> | 0.858<sub>(0.846,0.871)</sub> | 0.434<sub>(0.403,0.467)</sub> |
| Inpatient Mortality | 0.853<sub>(0.834,0.874)</sub> | 0.200<sub>(0.163,0.251)</sub> | 0.856<sub>(0.842,0.872)</sub> | 0.243<sub>(0.205,0.282)</sub> |

**Figure 2.**
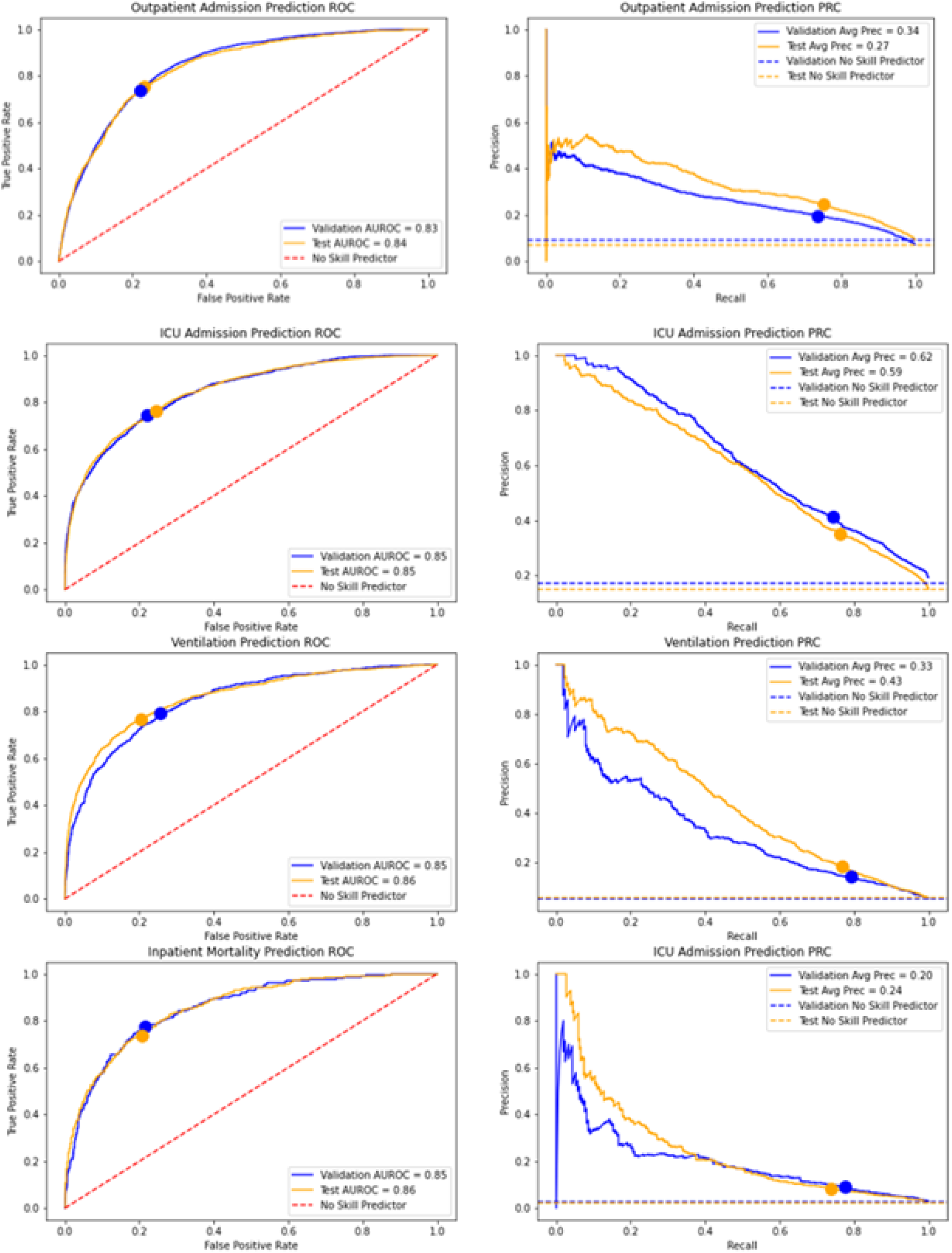
Classification performance metrics. ROC (left) and PRC (right) for the models considered (top to bottom): hospital admission, ICU admission, ventilation and inpatient mortality. Dashed lines represent no skill (random) predictions. Classification thresholds selected using Youden’s methods on only the validation set of each model are denoted as dots.

The operational point of each model, *i.e.*, the classification threshold, *τ*, over which the model will call a patient as being admitted to the hospital, or admitted to the ICU, ventilated and/or dying in an inpatient setting, were selected on the validation set using Youden’s method and highlighted in Figure 2. Table 4 shows summaries of the confusion matrix, *i.e.*, TPR (True Positive Rate, or sensitivity), TNR (True Negative Rate, or specificity), FPR (False Positive Rate), FNR (False Negative Rate), PPV (Positive Predictive Value, or precision), NPV (Negative Predictive Value) and Accuracy (overall agreement), for the test set along with the selected thresholds. See Table S2 in the Supplement for similar summaries on the validation set.

**Table 4.** Confusion matrix summaries on the test set. Thresholds, *τ*, for each model were selected using Youden’s method on the validation set. The reported metrics include: TPR (True Positive Rate, or sensitivity), TNR (True Negative Rate, or specificity), FPR (False Positive Rate), FNR (False Negative Rate), PPV (Positive Predictive Value, or precision), NPV (Negative Predictive Value) and Accuracy (overall agreement). Figures in parentheses are 95% CIs estimated via bootstrapping.

| Model | $\tau$ | TPR | TNR | FPR | FNR | PPV | NPV | Accuracy |
| --- | --- | --- | --- | --- | --- | --- | --- | --- |
| Admission | 0.101 | 73.5<sub>(72.0,74.9)</sub> | 78.1<sub>(77.8,78.5)</sub> | 21.8<sub>(21.5,22.2)</sub> | 26.5<sub>(25.1,28.0)</sub> | 19.6<sub>(18.9,20.3)</sub> | 97.6<sub>(97.4,97.6)</sub> | 77.8<sub>(77.5,78.2)</sub> |
| ICU Admission | 0.154 | 76.4<sub>(74.7,77.7)</sub> | 75.5<sub>(75.0,76.2)</sub> | 24.4<sub>(23.8,25.0)</sub> | 23.6<sub>(22.3,25.3)</sub> | 35.3<sub>(43.1,36.5)</sub> | 94.8<sub>(94.4,95.2)</sub> | 75.7<sub>(75.1,76.3)</sub> |
| Ventilation | 0.034 | 76.8<sub>(74.2,79.3)</sub> | 79.8<sub>(79.2,80.4)</sub> | 20.2<sub>(19.6,20.8)</sub> | 23.2<sub>(20.7,25.8)</sub> | 18.2<sub>(17.0,19.3)</sub> | 98.3<sub>(98.1,98.5)</sub> | 79.6<sub>(79.1,80.2)</sub> |
| Inpatient Mortality | 0.014 | 73.8<sub>(70.3,77.4)</sub> | 79.7<sub>(79.1,80.2)</sub> | 20.3<sub>(19.8,20.9)</sub> | 26.2<sub>(22.6,29.7)</sub> | 8.1<sub>(7.3,9.0)</sub> | 99.2<sub>(99.1,99.3)</sub> | 79.6<sub>(79.0,80.1)</sub> |

### Predictive Features

We identified the most important features for each model in terms of relative importance (see Section 2) during training. Figure 3 shows the top-10 features in order of relative importance for both the hospital admission model for the outpatient cohort (Figure 3 Top) and inpatient models (Figure 3 Middle). Examples of two of the identified features: max pulse (with missingness displayed separately) from the inpatient models and alcohol user from the outpatient model grouped by COVID-19 testing are presented in Figure 3 (Bottom).

**Figure 3.**
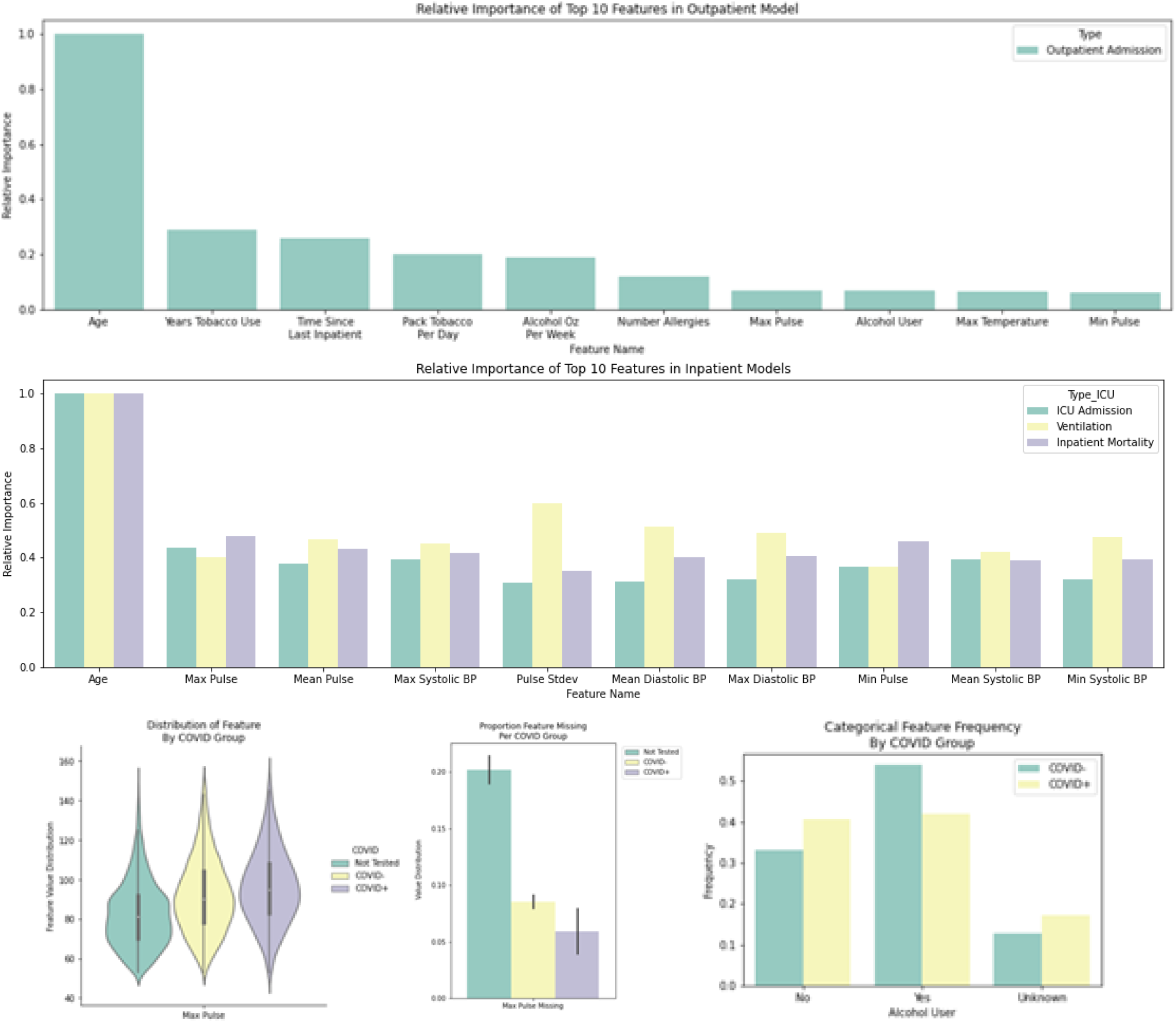
Predictor relative importance. The top-10 features by relative importance are shown for the outpatient model (Top) and inpatient models (Middle). Distribution summaries for two of the features with high relative importance stratified by COVID-19 testing (not tested, positive and negative) are also shown, *e.g.*, max pulse (Bottom left) and its missingness (Bottom Middle), and whether the individuals is an alcohol user (Bottom Right).

## 4 Discussion

From the cohort characteristics for the outpatient cohort in Table 1 we see that patient tested for COVID-19 with positive and negative results are different in terms of age, sex, race and all considered outcomes except for ventilation, for which we observe very similar ventilation rates. The COVID-19 positive patients (*N* = 3, 838) are characterized by a relatively younger population, lesser representation of White/Caucasian patients, nearly half the within-4-week admission rates and an inpatient mortality rate about five times higher relative to COVID-19 negative patients. Moreover, from the cohort characteristics for the inpatient cohort in Table 2 we see similar trends for the COVID-19 positive and negative or unknown COVID-19 results. However, age distributions between groups are more similar than that of the outpatient cohort. Interestingly, the inpatient mortality ratio between groups is relatively close to that of the outpatient cohort, yet ICU admissions and ventilation are much more prevalent in COVID-19 positive patients. Moreover, the COVID-19 positive patients experience longer LOS relative to patients that tested negative or that were not tested for COVID-19.

Model results in Figure 2 indicate good agreement between the validation and test results. Differences in the PRCs and their AP summaries can be explained in part by the differences in empirical prevalence (horizontal dashed lines) of the cases observed for each model, *e.g.*, ICU admissions. Further, the AUROC values for validation and test sets shown in Table 3 also support the performance similarity in both cohorts. The AP summaries are less similar, however, comparable and acceptable considering the low observed prevalences, particularly for the hospital admission model. Moreover, we see that the operational points (classification thresholds) selected via Youden’s method on the validation set (blue curves) result in well-matching the performance characteristics for both sets, validation and test. Summaries from the confusion matrix for the test set in Table 4 in general indicate good performance from all models in terms of TPR (76% average), TNR (77% average) and Accuracy (77% average). In terms of NPV and PPV, the very high NPV (97% average) suggest that that these models may be used for screening purposes while maintaining relatively high PPV values (22% average); considering the very low prevalences (8% average, with mortality being the lowest: 2.4%). Understandably, these thresholds were selected using Youden’s method as opposed to informing the threshold selection using criteria specific to the clinical practice, however, these can be readily addressed to suit the desired application without the need for re-training the models.

The key findings from the analysis of the most important features for the models considered and presented in Figure 3 are that: *i*) age is the most important feature (predictor), and *ii*) the most important features for the hospital admission and inpatient models are noticeably different. For instance, for the hospital admission model in the outpatient setting, social characteristics (alcohol and tobacco usage), the recency of the last inpatient encounter and allergy count, are the most predictive. However, for the inpatient model, summaries of physiological measurements, *e.g.*, pulse and blood pressure, are the most important. Importantly, we do not see comorbidities and laboratory results, included as inputs to the model (see Table S3 in the Supplement), playing an important role in the predictions from the model. This is interesting because it suggests that utilization and mortality can be predicted with expected performance as reported in Table 4 with fairly easy to obtain patient features.

The main difference between the proposed models and those previously proposed is that existing approaches^15–17^ mainly focus on mortality, whereas we also address hospital utilization, *e.g.*, hospital admissions within 4-weeks, ICU admissions and ventilation. Outcomes aside, most existing approaches use similar modeling strategies, *e.g.*, logistic regression, Cox proportional hazards, random forests and gradient boosting. In terms of features (predictors), approaches vary widely. However, there seems to be accumulating evidence that age and physiological summaries, such as vitals, are consistently predictive. Unlike^17^, we did not find comorbidities or laboratory results to be highly predictive (see Figure 3), though they were included as inputs to the model (see Table S3 in the Supplement). Though performance metrics across different studies are not directly comparable for a multitude of reasons, most of the existing approaches report AUCs within a similar range (AUROC: 0.85-0.95).

This study has several limitations. First, though the sample size of our study is large compared to other published works^16,17^, our cohort, largely pulled from the Durham-Raleigh area in North Carolina, may not be representative of the characteristics of the whole US population. Second, though results are validated chronologically, they are representative of a single health system (DUHS), thus the models developed here will need to be properly validated before being applied to other health systems. Third, from a data source perspective, features being extracted from an EHR, are limited to the availability, correctness and completeness of fields being considered (see Table S3 in the Supplement). Further, some patients may have incomplete features and outcomes in scenarios where they were seen at facilities outside DUHS. Fifth, though we count with the first five months worth of patient data since the beginning of the pandemic, it is likely that patient distribution will drift due to changing infection rates, treatment strategies, novel therapeutics and yet to be approved vaccines, which may change the performance characteristics of the model and the subset of most important predictors. Given these circumstances, we advocate for models that are refined (or retrained) periodically to take advantage of the sample size gains and to account for rapid changes in the population distributions.

As future work, we will aim to: *i*) expand the cohorts both in terms of sample size and features, possibly including other sources of data such as medical claims and social determinants of health; *ii*) pursue an external validation using cohorts from a different health system; and *iii*) further explore the differences in risk between COVID-19 positive and negative patients by leveraging approaches from the causal inference literature to better understand the outcome disparities after accounting for the observed heterogeneity of these two populations.

## 5 Conclusion

We demonstrated empirically that features aggregated from the EHR can be used for the stratification of patients tested for COVID-19 under different outcome scenarios. From a utilization perspective, patients can be ranked by likelihood of hospital admission up to 4 weeks in advance, ICU admission and the need for mechanical ventilation. From an outcomes perspective, patients can be ranked by likelihood of inpatient mortality. We also showed that the subset of EHR features is relatively small and dominated by Age and social indicators for the hospital admission model and by Age and physiological summaries for the inpatient endpoints (ICU admission, ventilation and mortality).

## Data Availability

The data used in this study is Protected Health Information and therefore cannot be shared publicly

## 6 Acknowledgements

The authors would like to thank the Duke Institute for Health Innovation (DIHI) for providing access to the COVID data and the scripts and resources for transforming it. This research was supported in part by NIH/NIBIB R01-EB025020.

## 7 Author Contributions

CD contributed with data acquisition, data curation, statistical modeling and analysis, draft writing, and revision and editing of the submitted manuscript. MG contributed with conceptualization, data acquisition, data curation, draft writing, and revision and editing of the submitted manuscript. MN contributed with data acquisition, and revision and editing of the submitted manuscript. RH contributed with conceptualization, supervision, draft writing, and revision and editing of the submitted manuscript

## 8 Additional Information

### Ethics Committee Approval

An exemption from institutional review board approval was determined by the DUHS Institutional Review Board as part of an active epidemiological investigation with no (current) medical intervention.

### SARS-CoV-2 Tests

Specific names indicating a COVID-19 test within EPIC’s Clarity database were identified with clinical guidance. The list contains tests of the following types from different distributors: SARS-COV-2 real time PCR tests, SARS-CoV-2 nucleic acid amplification tests, pan-SARS and SARS-CoV-2 specific RNA tests, and point of care SARS-CoV-2 rapid tests.

### Features used for Modeling

The complete list of features used as inputs to the models is summarized in Table S3 in the Supplement.

