## Supplemental Information for "Predicting Hospital Utilization and Inpatient Mortality of Patients Tested for COVID-19"

[1]**Chief complaints:** Abdominal pain, abnormal lab, abnormal radiology image, abscess, alcohol intoxication, alcohol problem, altered mental status, arm swelling, back pain, back pain - lumbar area, bloated, blood infection, blood pressure check, bradycardia, cardiac arrest, chest pain, chest pain - non-traumatic, chills, constipation, contractions, cough, dehydration, dialysis - needs, diarrhea, dizziness, drug overdose, dysphagia, dysuria, emesis, epistaxis, extremity weakness, eye problem, facial swelling, fall, fatigue, fever, flank pain, flu-like symptoms, foot injury, generalized body aches, generalized weakness, groin swelling, gunshot wound, hand pain, headache, hematemesis, hematuria, hemoptysis, hip injury, hip pain, hyperglycemia, hypertension, hypoglycemia, hypotension, irregular heartbeat, jaundice, knee pain, laboring, leg injury, leg pain, leg swelling, loss of consciousness, LVAD complication, melena, motor vehicle crash, motorcycle crash, nausea, nausea vomiting diarrhea, neck pain, neurologic problem, non-stress test, numbness, oncology and fever, other, pain, palpitations, post-op problem, psychiatric evaluation, rash, rectal bleeding, respiratory distress, rupture of membranes, scheduled C-section, scheduled induction, seizures, shortness of breath, shoulder pain, sickle cell pain crisis, sore throat, stroke like symptoms, tachycardia, transplant and fever, trauma, urinary frequency, urinary retention, vaginal bleeding, vascular access problem, withdrawal, wound check, wound infection.

[2]**Laboratory tests:** BUN, CKMB, CRP, ESR, INR, LDH, arterial PCO2, venous PCO2, arterial PO2, venous PO2, PTH, albumin, ALT, ammonia, anion gap, APTT, AST, band count, arterial bicarbonate, venous bicarbonate, bilirubin, BNP, total arterial CO2, total venous CO2, creatine kinase, creatinine, D-dimer, fibrinogen, glucose, HCT, hemoglobin A1C, ionized calcium, lactate, lipase, magnesium, arterial pH, venous pH, platelet, potassium, sodium, troponin I, HS troponin T, WBC.

[3]**Medication Administration Record (MAR):** Acetaminophen items associated with admission from the emergency department, acetaminophen items associated with discharge from the emergency department, albuterol, alprazolam, amiodarone, amoxicillin, aspirin, atorvastatin, atropine, azithromycin, budesonide, calcium gluconate, carvedilol, cefepime, clindamycin, clonazepam, clonidine, dexamethasone, dextrose items associated with admission from the emergency department, dextrose items associated with discharge from the emergency department, diazepam, dicyclomine, diltiazem, diphenhydramine, divalproex, DTaP, enoxaparin, epinephrine, eptifibatide, famotidine, furosemide, GI cocktail, haloperidol, heparin items associated with admission from the emergency department, heparin items associated with discharge from the emergency department, hydrocodone, hydromorphone items associated with discharge from the emergency department, ibuprofen, individual MAR items associated with ICU admission from the emergency department, individual MAR items associated with admission from the emergency department, individual MAR items associated with discharge from the emergency department, insulin, iopamidol, ipratropium, KCl associated with admission from the emergency department, KCl items associated with discharge from the emergency department, ketorolac, levetiracetam, levothyroxine, lidocaine, lisinopril, lorazepam, lactated ringer, metformin, metoclopramide, metoprolol, metronidazole, midazolam, NaCl items associated with admission from the emergency department, NaCl items associated with discharge from the emergency department, naproxen, nicotine, norepinephrine, olanzapine, ondansetron, oxycodone, pantoprazole, prochloperazine, promethazine, quetiapine, sennosides, trazodone, valproate, vancomycin.

[4]**Orders:** Abdomen x-ray, arterial blood gas, ankle x-ray, basic metabolic panel, catheter lab, complete blood count, drug toxicology labs, ECG, echocardiogram, elbow x-ray, foot x-ray, forearm x-ray, supplemental oxygen, hand/finger x-ray, head/neck CT scan, hip/pelvis x-ray, humerus x-ray, individual orders associated with admission from the emergency department, individual orders associated with discharge from the emergency department, knee x-ray, portable knee x-ray, pregnancy test, ribs x-ray, shoulder x-ray, skin test, spine x-ray, tibia/fibula x-ray, urinary analysis, ultrasound, venous blood gas, ventilation, wrist x-ray.

[5]**Vitals:** Diastolic blood pressure, height, oxygen flow rate, pulse, pulse oximetry, respiratory rate, systolic blood pressure, temperature, weight.

**Table S1.** Range of hyperparameter values swept by RandomizedSearchCV for the LightGBM models of admission within 4 weeks of outpatient COVID-19 test, ICU admission, ventilation, and inpatient mortality.

| Parameter Name | LightGBM Name | Values |
|---|---|---|
| Max Depth | max_depth | [-1, 4, 6, 12] |
| Number of Estimators | n_estimators | [500, 1200, 1500, 2500, 5000] |
| Number of Leaves | num_leaves | [5, 25, 50, 60, 75, 100] |
| Minimum Data Per Leaf | min_child_samples | [100, 250, 500, 750, 1000] |
| Minimum Instance Weight Per Leaf | min_child_weight | [0.1, 1, 10, 20, 100, 1000] |
| Subsample Ratio of Columns Per Tree | colsample_bytree | [0.1, 0.3, 0.5, 0.7, 0.9] |
| L1 Regularization | reg_alpha | [0, 0.1, 1, 5, 10, 50] |
| L2 Regularization | reg_lambda | [0, 0.1, 1, 5, 10, 50] |
| Learning Rate | learning_rate | [0.001, 0.005, 0.01] |

**Table S2.** Confusion matrix summaries on the validation set. Thresholds, $\tau$, for each model were selected using Youden's method on the validation set. The reported metrics include: TPR (True Positive Rate, or sensitivity), TNR (True Negative Rate, or specificity), FPR (False Positive Rate), FNR (False Negative Rate), PPV (Positive Predictive Value, or precision), NPV (Negative Predictive Value) and Accuracy (overall agreement). Figures in parentheses are 95% CIs estimated via bootstrapping.

| Model | $\tau$ | TPR | TNR | FPR | FNR | PPV | NPV | Accuracy |
|---|---|---|---|---|---|---|---|---|
| Admission | 0.101 | $75.2_{(73.0,77.5)}$ | $76.9_{(76.1,77.7)}$ | $23.1_{(22.3,23.8)}$ | $24.8_{(22.5,27.0)}$ | $24.5_{(23.2,25.8)}$ | $96.9_{(96.6,97.2)}$ | $76.7_{(76.1,77.4)}$ |
| ICU Admission | 0.154 | $74.4_{(72.5,76.3)}$ | $77.9_{(76.9,78.8)}$ | $22.1_{(21.2,23.1)}$ | $25.6_{(23.7,27.5)}$ | $41.1_{(39.4,43.0)}$ | $93.6_{(93.1,94.1)}$ | $77.3_{(76.5,78.0)}$ |
| Ventilation | 0.034 | $79.0_{(75.4,82.8)}$ | $74.4_{(73.6,75.3)}$ | $25.5_{(24.7,26.4)}$ | $21.0_{(17.2,24.6)}$ | $14.1_{(13.0,15.3)}$ | $98.5_{(98.3,98.8)}$ | $74.7_{(73.8,75.5)}$ |
| Inpatient Mortality | 0.014 | $77.0_{(72.3,81.9)}$ | $78.6_{(77.8,79.4)}$ | $21.4_{(20.6,22.2)}$ | $23.0_{(18.1,27.7)}$ | $9.1_{(8.0,10.4)}$ | $99.2_{(99.0,99.4)}$ | $78.5_{(77.8,79.3)}$ |

**Table S3.** Description of all data types and elements used for outpatient modeling (predicting admission within 4 weeks of an outpatient COVID-19 test) and inpatient modeling (ICU admission, ventilation, and inpatient mortality).

| Data Category | Inpatient Elements | Outpatient Elements |
|---|---|---|
| Demographic | Age | Age<br>Race<br>Sex |
| Encounter Specific | Time since last inpatient encounter<br>Number of prior encounters | Time since last inpatient encounter<br>Number of prior encounters |
| Allergies | NA | Number of active allergies |
| Social | NA | Alcohol use (yes/no, ounces per week)<br>Tobacco use (yes/no, years used, packs per day) |
| Prior Procedures | See CPT to Single CCS Category Crosswalk | See CPT to Single CCS Category Crosswalk |
| Problem List | See ICD10 to Single CCS Category Crosswalk | See ICD10 to Single CCS Category Crosswalk |
| Comorbidities (ICD 10 Diagnoses) | See ICD10 to Single CCS Category Crosswalk | NA |
| Chief Complaint | Full list above[1] | NA |
| Laboratory Tests | Indicator for collection of lab and lab value<br>Full list above[2] | NA |
| MAR | Indicator for administration of medication<br>Full list above[3] | NA |
| Orders | Indicate for order placed<br>Full list above[4] | NA |
| Vitals | For each item, calculated max, min, mean, standard deviation, and number of times recorded<br>Full list above[5] | For each item, calculated max, min, mean, standard deviation, and number of times recorded |

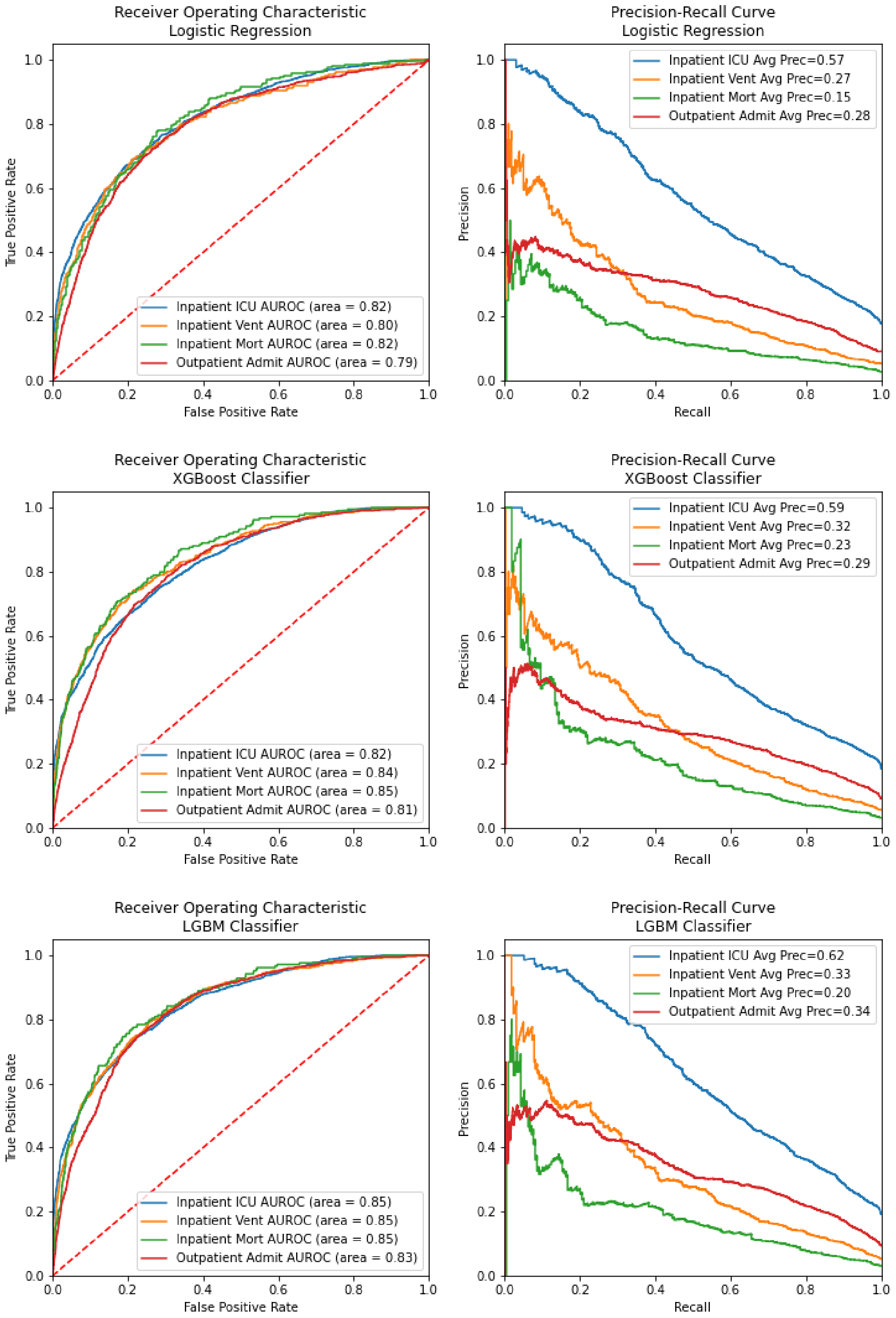


**Figure S1.** Performance of initial Logistic Regression, XGBoost Classifier, and LightGBM classifier models, built on the train sets with default hyperparameters and evaluated on the validation sets.

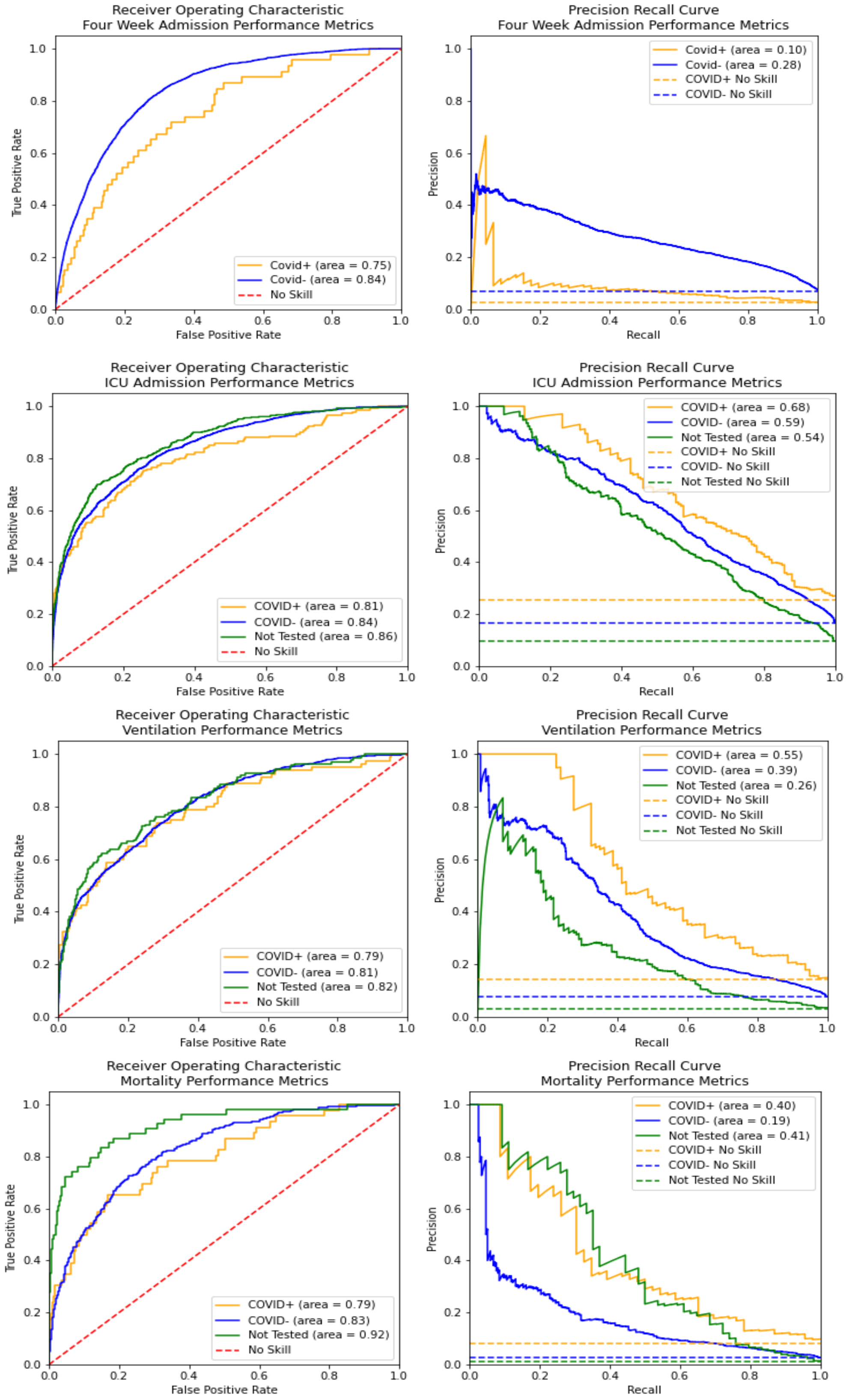


**Figure S2.** Classification performance metrics. ROC (left) and PRC (right) for the models considered (top to bottom): hospital admission within 4 weeks, ICU admission, ventilation and inpatient mortality grouped by COVID-19 testing status (positive, negative and not tested). Dashed lines represent no skill (random) predictions.
